# Risk Factors Prediction, Clinical Outcomes and Mortality of COVID-19 Patients

**DOI:** 10.1101/2020.07.07.20148569

**Authors:** Roohallah Alizadehsani, Zahra Alizadeh Sani, Mohaddeseh Behjati, Zahra Roshanzamir, Sadiq Hussain, Niloofar Abedini, Fereshteh Hasanzadeh, Abbas Khosravi, Afshin Shoeibi, Mohamad Roshanzamir, Pardis Moradnejad, Saeid Nahavandi, Fahime Khozeimeh, Assef Zare, Maryam Panahiazar, U. Rajendra Acharya, Sheikh Mohammed Shariful Islam

## Abstract

**Background:** Preventing communicable diseases requires understanding the spread, epidemiology, clinical features, progression, and prognosis of the disease. Early identification of risk factors and clinical outcomes might help to identify critically ill patients, provide proper treatment and prevent mortality.

**Methods:** We conducted a prospective study in patients with flu-like symptoms referred to the imaging department of a tertiary hospital in IRAN between 3 March 2020 and 8 April 2020. Patients with COVID- 19 were followed up to check their health condition after two months. The categorical data between groups were analyzed by Fisher’s exact test and continuous data by Wilcoxon Rank-Sum Test.

**Findings:** 319 patients (mean age 45.48±18.50 years, 177 women) were enrolled. Fever, dyspnea, weakness, shivering, C-reactive protein (CRP), fatigue, dry cough, anorexia, anosmia, ageusia, dizziness, sweating and age were the most important symptoms of COVID-19 infection. Traveling in past three months, asthma, taking corticosteroids, liver disease, rheumatological disease, cough with sputum, eczema, conjunctivitis, tobacco use, and chest pain did not have any relationship with COVID-19.

Research in context

Evidence before this study
We searched Google scholar, PUBMED and Scopus for articles that investigated the recent epidemic of COVID-19, especially those that investigate effective risk factors. We found that there is not enough research in this field, especially the risk factor that is effective in finding the rate of mortality of this disease.

Added value of this study
We determined some of the most important effective risk factors on prediction, clinical outcome and mortality rate of COVID-19 infection. To the best of our knowledge, some of these risk factors are investigated in this work for the first time. Our findings could provide good insight into the early prediction of the disease, its clinical outcomes, and suggest a cost-effective method for mortality prediction.

Implication of all the available evidence
COVID-19 can transmit human-to-human and lead to severe symptoms and high mortality. Early prediction of this disease and the risk of mortality can help the physicians to better manage this worldwide health problem.

**Interpretation:** Finding clinical symptoms for early diagnosis of COVID-19 is a critical part of prevention. These symptoms can help in the assessment of disease progression. To the best of our knowledge, some of the effective features on the mortality due to COVID-19 are investigated for the first time in this research.

**Funding:** None

## Introduction

In Wuhan, Hubei Province, China, since December 2019, a cohort of patients suffered from acute respiratory disease with unknown etiology [1]. The first cases exhibited a link to the Huanan wholesale seafood market. By investigating the throat swab sample of patients, the Chinese Center for Disease Control and Prevention (CDC) detected a new coronavirus, that was previously known as severe acute respiratory syndrome coronavirus 2 (SARS-CoV-2, formerly dubbed as 2019-nCoV) by the International Committee on Taxonomy of Viruses (ICTV) [2]. Human ACE2 molecules have a strong interaction with the receptor-binding domain (RBD) of SARS-CoV-2 Spike protein. This yields a high infectious competence of the virus to human respiratory epithelial cells. The virus causes fatigue, cough, fever and severe or mild respiratory impediments. SARS-CoV-2 is termed as Coronavirus Disease 2019 (COVID-19), with which most of the patients exhibited mild symptoms [3]. But a fraction of the critical patients developed acute respiratory failure, septic shock and other grim complications including acute respiratory distress syndrome (ARDS), and multiple organ dysfunction syndromes (MODS) that may lead to fatal outcomes [4]. COVID- 19 has been recognised as a Class B respiratory contagious ailment by the China Health Committee.

The WHO, on March 12th, 2020, declared COVID-19 as a global pandemic [5]. Several measures were implemented to restrain the outbreak of the disease by different governments across the globe. These measures include restrictive face-to-face communications via obligatory ‘social distancing’ and closure of public education and recreation sites such as parks, schools, colleges and Universities. Majority of the world’s population have gone through an unprecedented experience by following stern observance to the new measures. Elderly persons, along with some ailments such as acute kidney injury, diabetes mellitus, cardiovascular diseases, cancer and hypertension are at higher risk of mortality or may have more critical of COVID-19 [4]. 13.8–19.1% reported COVID-19 patients in Wuhan, China became ailments critically. An astonishing fatality rate of 61.5% was recorded in the latest reports that increased significantly with age and for patients with comorbidities[6]. This has led to the scarcity of intensive care facilities in the hospitals due to the exponential increase in the number of cases and also put enormous pressure on medical staff and services. It is unfortunate not to have any prognostic biomarker to identify patients that may need urgent medical care and associated fatality rate. Besides, although there is a rapid increase in COVID-19 cases, the information concerning the clinical symptoms (features) is inadequate. Liu et al. [7] compared the clinical characteristics of elderly patients of COVID-19 with middle-aged and young patients. The common symptoms were fever, sputum and cough. The proportion of multiple lobe involvement and pneumonia severity index (PSI) score (p<0.001) of the old patient’s group was elevated than the middle and young aged cohort.

Early detection of disease helps the clinicians to provide necessary, timely treatment. Zheng et al. [8] explored the clinical and epidemiological characteristics of coronavirus. They used the treatment, radiological, laboratory, clinical, demographic and epidemiological data of 99 confirmed COVID-19 patients in China. They identified fever, fatigue and dry cough as common symptoms. The median age of recorded patients is 49 years, 41% had the underlying disease, 49% came in close contact with the COVID- 19 affected patients, and 42% had lived in or travelled to Wuhan. Lower CD8 and CD4 counts, smaller white blood cells, lymphocytes and neutrophils; higher brain natriuretic peptide levels; higher levels of myocardial damage and higher C-reactive protein levels may be used for the early recognition of severely ill patients of the disease.

Previous studies have shown that COVID-19 patients present with different signs and symptoms. Abnormal liver function is observed in some of the COVID-19 patients. Fan et al. [9] studied the features related to COVID-19 associated liver damage for providing the treatment. The study included 75 male and 73 female patients from China with a mean age of 50 years. The enhanced levels of total bilirubin, alkaline phosphatase, gamma-glutamyltransferase and aspartate and alanine aminotransferase are considered as abnormal liver function. The patients with abnormal liver function are treated with lopinavir/ritonavir drug after hospital admission. Such patients had more extended hospital stay than patients with normal liver function.

Case-fatality rates and confirmed cases of COVID-19 are different among countries. One probable reason may be universal bacilli Calmette-Guérin (BCG) vaccine coverage varies from country to country. Hamiel et al. [10] reviewed 72,060 test results among the COVID-19 affected patients from Israel. They did not find a statistical difference between the positive test results in the unvaccinated group and BCG vaccinated cohort. Their study could not find any protective effect on the BCG vaccinated adults against the pandemic.

Heart transplant (HT) patients may have higher risk levels from COVID-19 disease due to clinically significant immunosuppression and several other comorbidities. Latif et al. [11] studied the treatment, outcomes and characteristics of patients having COVID-19 with HT. Their study recorded the fatality rate of 25% in patients of HT related to COVID-19 disease. Wu et al. [12] exploited the outcomes and the clinical characteristics in COVID-19 infected patients who died or had acute respiratory distress syndrome (ARDS). They extracted the risk factors related to ARDS development and death as coagulation dysfunction, neutrophilia and older age by using bivariate Cox regression. They concluded that due to less immune response, former age patients have a higher risk of ARDS development and death. Rothe et al. [13] examined the transmission of COVID-19 disease from asymptomatic contact in Germany. They concluded that asymptomatic patients were potential carriers of COVID-19 infection, and there was an urgent need for the examination of transmission dynamics of a pandemic.

Oxley et al. [14] investigated five patients of large-vessel stroke who were diagnosed with COVID-19 in a New York City hospital with a 5% prevalence of stroke among COVID-19 patients. Reluctance to present to the hospital, isolation, and social distancing were causes of the poor outcome. They asserted the need for further study of the association between COVID-19 in young patients and large-vessel stroke. First COVID- 19 case in Iceland was recorded in the last part of February, 2020. Gudbjartsson et al. [15] carried out screening with random samples from 2283 subjects, invited samples from 10,797 persons and out of which 643 were tested positive. Most of the tested positive persons had an international travel history at the early stage of the study. The haplotypes of the COVID-19 changed over time and were found to be diverse.

Type 2 diabetes (T2D) has been identified as prime comorbidity of COVID-19. It is uncertain about the effect of blood glucose (BG) control on the medical attention or mortality in patients with T2D as well as COVID-19. Zhu et al. [16] conducted a multi-centre, retrospective study of 7,337 patients in China, of which 952 already had Type 2 diabetes (T2D) disease. They observed that T2D were more prone to multiple organ injury, mortality and need more medical attention than non-diabetic ones. Early, fast and accurate clinical assessment of the COVID-19 severity level is crucial to support healthcare planning and decision making. Yan et al. [4] collected 485 COVID-19 patients’ blood samples from Wuhan, China, to detect critical biomarkers of the disease. Their machine learning method picked high-sensitivity C-reactive protein (hs-CRP), lymphocyte and lactic dehydrogenase (LDH) as their potential biomarkers that achieved 90% accuracy in mortality prediction. Some of the features that seem to have essential effects on COVID-19 mortality rate were not investigated in those papers. In this research, we analysed the additional risk factors of COVID-19 in Iran. To the best of our knowledge, there are a few studies available in this field, especially in Middle East countries.

### Method and material

This study was prospectively performed from 3 March 2020 to 8 April 2020 at the imaging department of OMID hospital, Tehran, IRAN. In this study, 319 patients were included with flu-like symptoms during the COVID-19 virus pandemic. All clinical data, including general information, epidemiological and medical history, symptoms, signs, epidemiological and clinical characteristics of patients, were included in this study. Totally 32 features (symptoms) were selected on consultation with four infection disease specialists. Then these features were included in the questionnaire of patients. This study was also approved by the local ethical committee of the university. Our patients were informed about the study aims, and written consent was obtained before the inclusion of this study.

For patients with symptoms, lung computed tomography (CT) has been performed as a noninvasive test for lung situation assessment. Unlike RT-PCR, which requires specific laboratory environments, CT-scan was used to provide a faster diagnosis of lung diseases. All the suspected patients underwent a thin-slice high-resolution multi-slice spiral CT scan in a supine position, and high-resolution computed tomography (HRCT) images of all patients were reviewed by a radiologist with more than 14 years of experience in chest imaging. This way, COVID-19 was diagnosed in suspicious cases. We have followed-up patients for two months after participation to determine their vital status.

### Statistical analysis

Data are presented as means (± standard deviations). We analyzed the features using Matlab 2016a software. Fisher’s exact test [17] and Wilcoxon Rank-Sum Test [18] are used for categorical and continuous data respectively, to specify differences between the two groups. Statistical significance is set at p ≤ 0.05.

## Results

A total 319 patients (mean age 45.48±18.50 years, 177 women) were recruited. In the patients with COVID- 19, one had leukemia, one advanced thyroid, and one bone marrow cancer and unfortunately the patients with leukemia and bone marrow cancer died. Meanwhile, two cases had a stroke and one of them was cured One patient with a history of Tuberculosis died. Two cases had kidney disease and both of them died.

Our data showed significant difference between healthy and COVID-19 cases with regard to the symptoms like fever (P-value=**1.99E-12**), dyspnea (P-value= **2.99E-11**), weakness (P-value=**3.16E-11**), shivering (P-value=**1.01E-09**), fatigue (P-value=**6.60E-09**) and dry cough (P-value=**9.53E-09**). Indeed, symptoms such as anorexia (P-value=**1.68E-08**), anosmia (P-value=**5.46E-08**), ageusia (P-value=**1.19E-07**), dizziness (P-value=**2.10E-05**) and sweating (P-value=**2.15E-05)** showed significant difference between healthy vs. COVID-19 cases as well. All of these symptoms are more prevalent in COVID-19 affected cases than healthy subjects. While considering symptoms such as chest pain, sore throat and cough with sputum, there is no significant difference between healthy vs. COVID-19 cases (P-value of 0.411, 0.666 and 1, respectively). The significantly higher *mean* age is seen in COVID-19 cases (52.02 ± 17.63-year-old) versus healthy subjects (44.13 ± 16.17 years old) (P-value=**1.54E-04**). Abnormal CRP is significantly more in COVID-19 affected cases compared to healthy subjects (P-value=**1.59E-09**). The O- BG (P-value=0.0066) is found to be lower for COVID-19 cases compared to normal subjects. There is no significant difference between healthy and COVID-19 cases regarding AB-, A-, A+, B+, AB+, B- and O+ BG carriers with P-values of 0.641, 0.6251, 0.6561, 0.7291, 0.8044, 1 and 1, respectively. Past history of BCG vaccination did not show any significant difference between COVID-19 and. healthy subjects (P-value= 0.1057). There is no significant difference between healthy and COVID-19 cases regarding DM (P-value= 0.269), immune deficiency (P=0.302), HEM (P-value= 0.377), rheumatologic diseases (P-value=0.377), corticosteroid therapy (P-value= 0.385), tobacco use (P-value= 0.38) and gender (P-value= 0.411). History of travel within the past *three* months showed no significant difference between the two classes (P-value= 0.546). We observed no significant difference between healthy and COVID-19 cases with regard to asthma (P-value= 0.715), liver disease (P-value= 0.746), cancer (P-value= 0.754), heart disease (P-value= 1), kidney disease (P-value= 1) and organ transplant (P-value= 1). The summary of this data is shown in Table 1. Features with a significant relationship with COVID-19 are illustrated in Figure 1. Meanwhile, the age distribution and the number of different blood type groups of COVID-19 patients are shown in Figure 2. Figure 2(a) shows that the groups with the highest infection risk are people in the age between 25 and 55. They are active labor force of society and have higher interaction with others. It is clear from Figure 2(b) that the negative blood groups are infected extremely less than positive groups. In positive blood groups, O+BG, A+BG, B+BG and AB+BG are more infected, respectively.

**Table 1.** Clinical characteristics of COVID-19 patients.

| Feature | Covid-19 | Healthy | P-Value |
| --- | --- | --- | --- |
| 'Fever' | 50 | 15 | **1.99E-12** |
| 'dyspnea' | 61 | 29 | **2.99E-11** |
| 'Weakness' | 34 | 5 | **3.16E-11** |
| ' Shivering ' | 32 | 6 | **1.01E-09** |
| 'CRP' | 84 | 66 | **1.59E-09** |
| 'Fatigue' | 38 | 12 | **6.60E-09** |
| 'Dry Cough' | 55 | 29 | **9.53E-09** |
| 'Anorexia' | 26 | 4 | **1.68E-08** |
| 'Anosmia' | 33 | 10 | **5.46E-08** |
| 'Ageusia' | 31 | 9 | **1.19E-07** |
| ' Dizziness ' | 11 | 0 | **2.10E-05** |
| 'Sweating' | 15 | 2 | **2.15E-05** |
| 'Age' | 52.02 ± 17.63 | 44.13 ± 16.17 | **1.54E-04** |
| 'Blood Type' |  |  |  |
| O- | 1 | 15 | **0.0066** |
| AB- | 2 | 2 | 0.641 |
| A- | 1 | 4 | 0.6524 |
| A+ | 24 | 34 | 0.6561 |
| B+ | 16 | 23 | 0.7291 |
| AB+ | 6 | 12 | 0.8044 |
| B- | 1 | 2 | 1 |
| O+ | 35 | 55 | 1 |
| BCG Vaccine | 88 | 158 | 0.1057 |
| 'Diabetes' | 10 | 24 | 0.269539 |
| 'Immunodeficiency' | 0 | 4 | 0.302357 |
| 'HEM' | 3 | 2 | 0.37745 |
| 'Rheumatological Disease' | 3 | 2 | 0.37745 |
| 'Corticosteroids' | 1 | 0 | 0.38558 |
| 'Tobacco' | 1 | 0 | 0.38558 |
| 'Gender' |  |  | 0.4116 |
| Male | 62 | 80 |  |
| Female | 61 | 116 |  |
| 'Chest Pain' | 1 | 5 | 0.411622 |
| 'Traveling In Past 3 Months Ago' | 6 | 6 | 0.546959 |
| 'Sore Throat' | 8 | 16 | 0.666779 |
| 'Asthma' | 2 | 6 | 0.715367 |
| 'Liver Disease' | 3 | 7 | 0.746248 |
| 'Cancer' | 5 | 6 | 0.754812 |
| 'Heart Disease' | 12 | 20 | 1 |
| 'kidney Disease' | 5 | 8 | 1 |
| 'transplant' | 0 | 1 | 1 |
| 'Cough With Sputum' | 2 | 4 | 1 |

**Figure 1.**
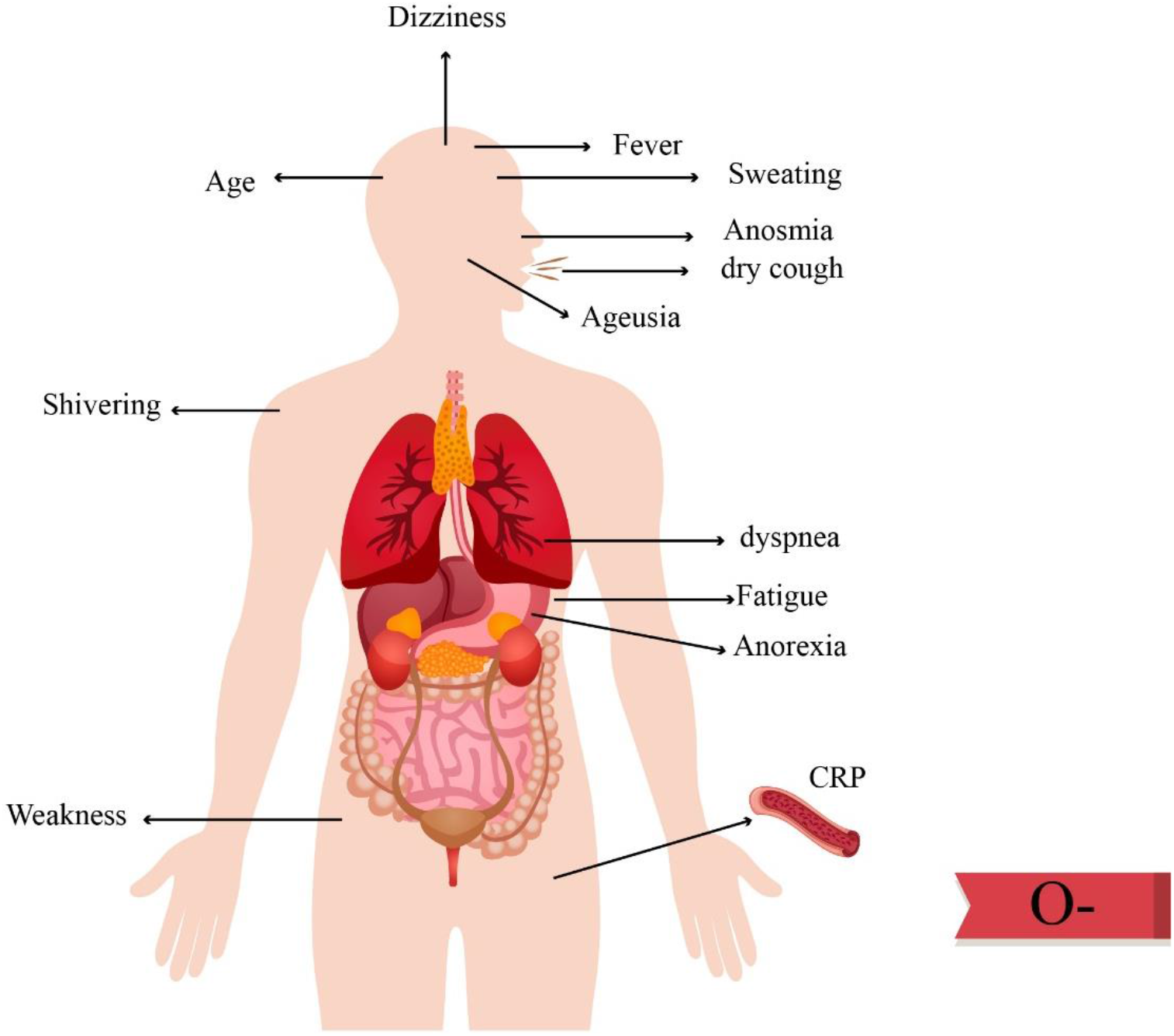
Features with a significant relationship with COVID-19.

**Figure 2.**
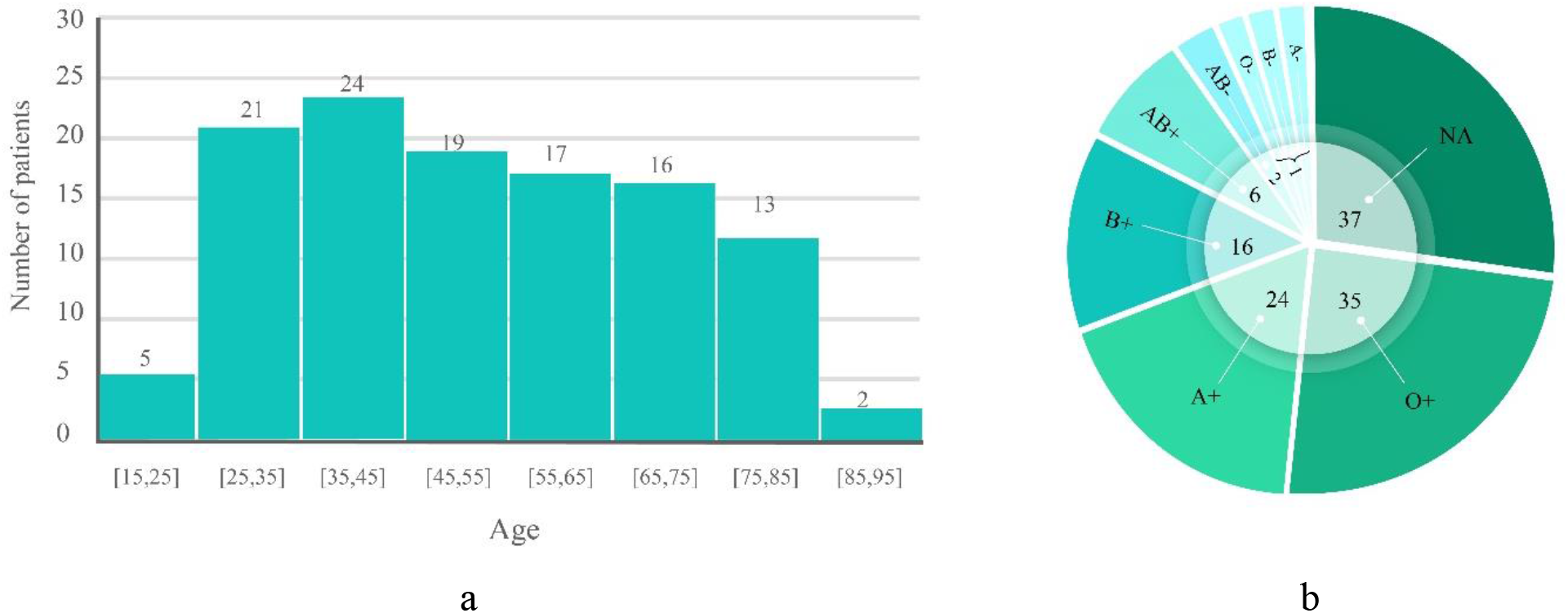
a) The age distribution of COVID-19 patients. b) Blood Types of COVID-19 Patients.

We observed a significant association between older age (P-value=**2.82E-05**), history of heart disease (P-value=**0.00654**), and history of cancer (P-value=**0.012863**) with COVID-19 mortality compared to healthy subjects. Carriers of O+BG showed protective features against COVID-19 with mortality (P-value= **0.0057**). Regarding symptoms, anosmia (P-value=**0.010612**), dry cough (P-value=**0.011324**), ageusia (P-value=**0.011741**), fever (P-value=**0.024933**) and anorexia (P-value=**0.038981**) are significantly related to COVID-19 with mortality compared to healthy subjects. Other features did not show a significant relationship with COVID-19 with mortality. The summary of this data is shown in Table 2. Features with a significant relationship with mortality in COVID-19 are shown in Figure 3. The age distribution of died patients because of COVID-19 is illustrated in Figure 4. According to this figure, although most in the infected people are in the range of 25 and 55, the mortality rate is so low. There is no mortality in the age between 40 and 60. But it does not mean that the youth are completely immune from death. There are 2 cases died in the range of 30 and 40. Although the infection rate is not high between older people, their mortality rate is high.

**Table 2.**
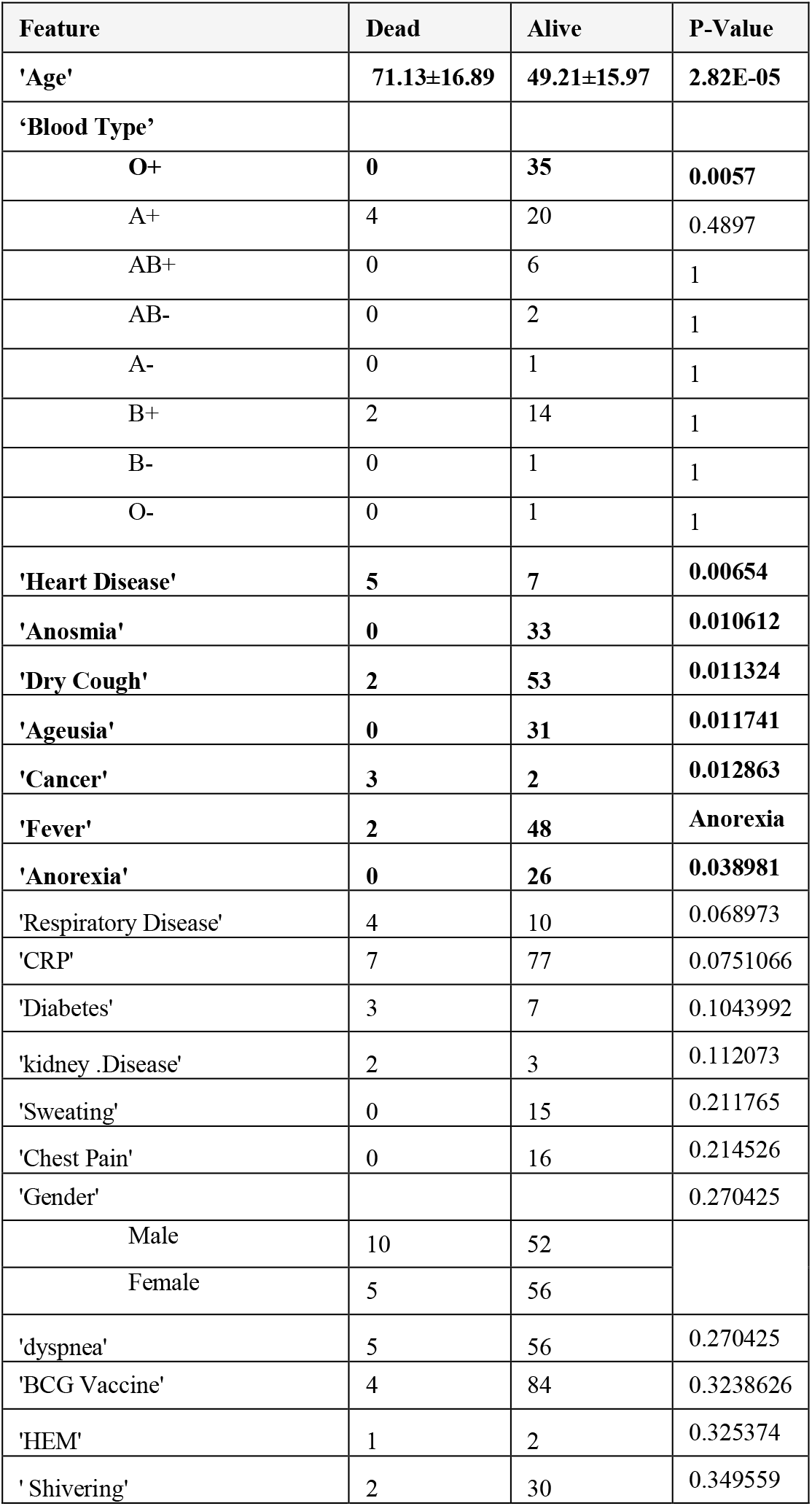
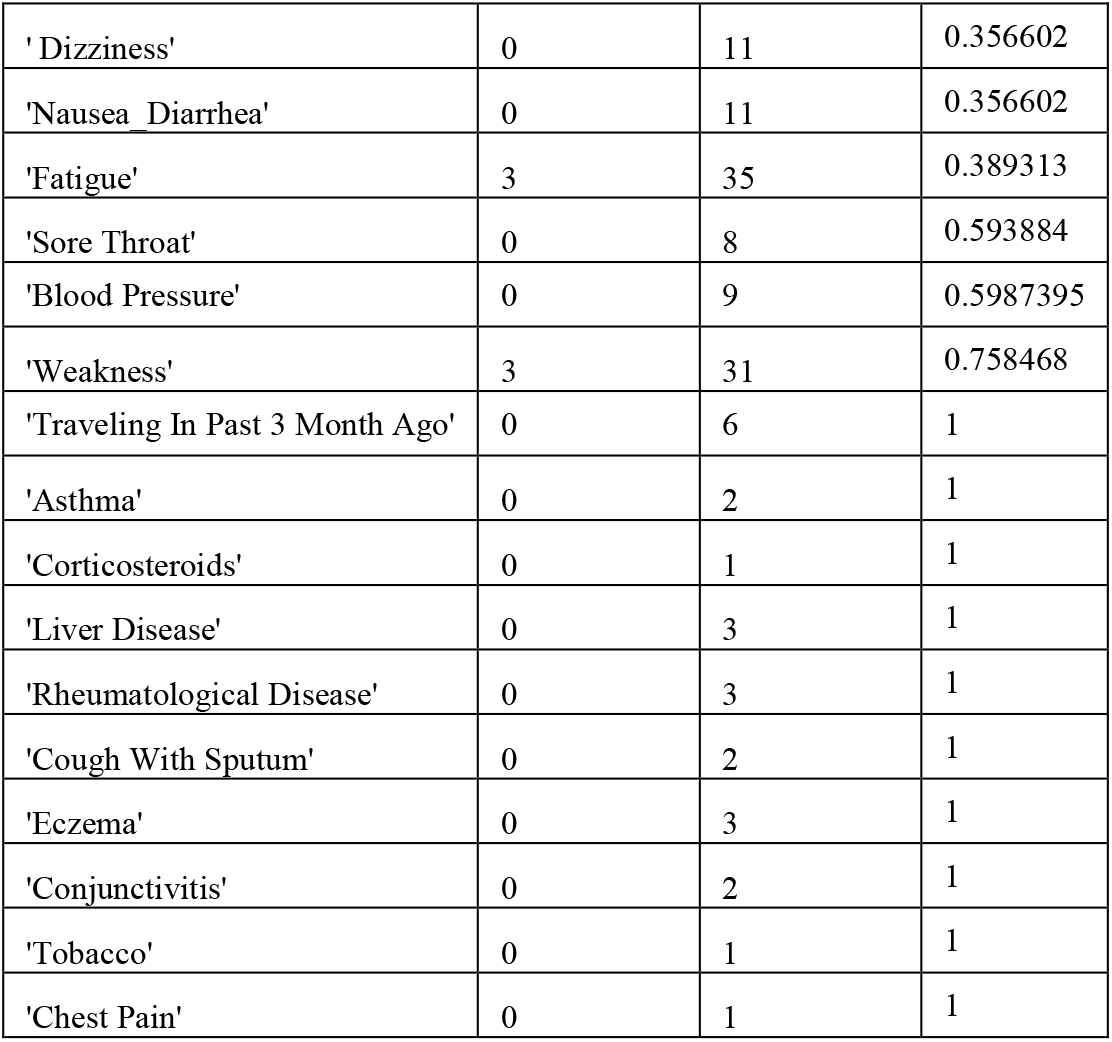
Clinical characteristics of COVID-19 patients with mortality.

**Figure 3.**
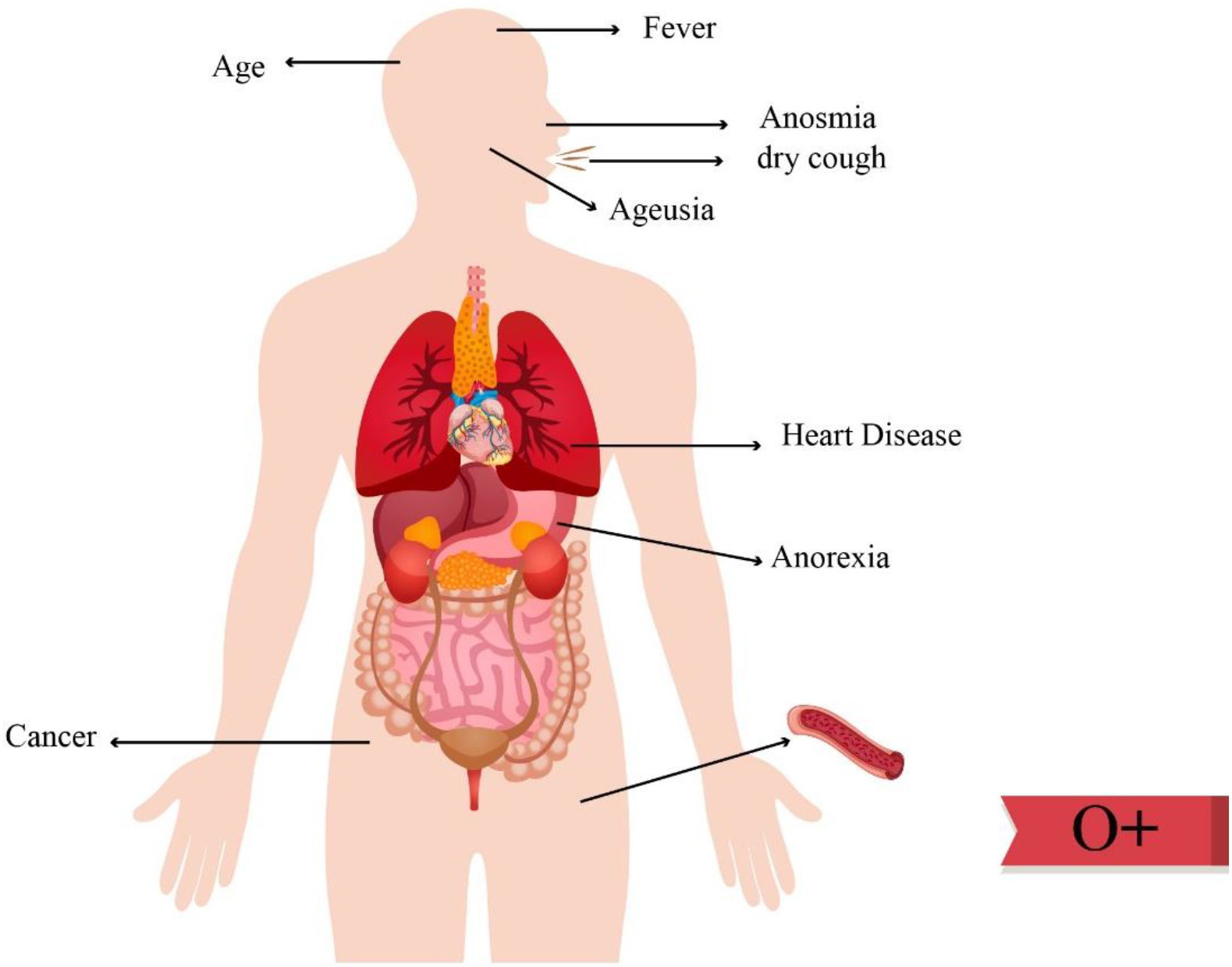
Features with a significant relationship with mortality in COVID-19.

**Figure 4.**
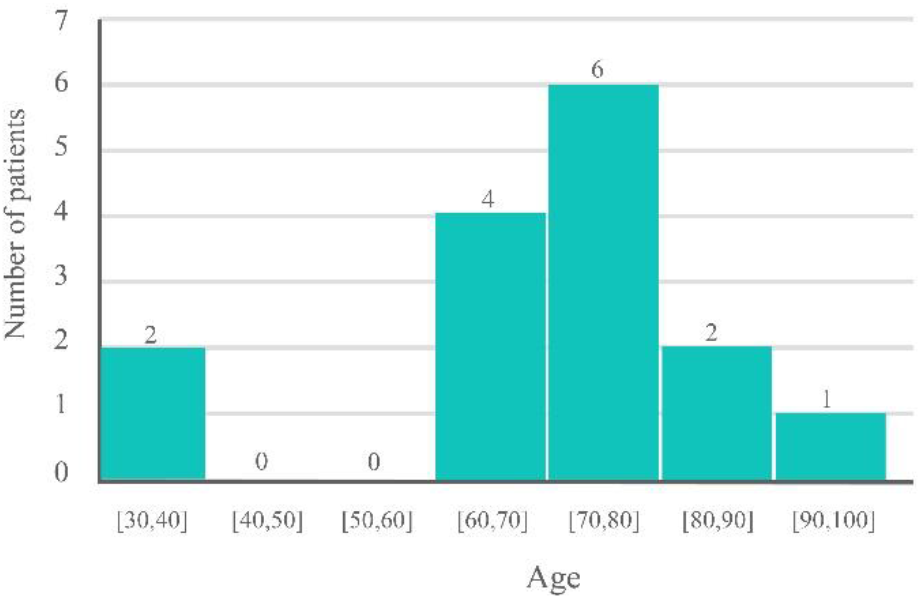
Age distribution of patients died because of COVID-19.

## Discussion

The main findings of our study are the significant association between symptoms such as fever, dyspnea, weakness, shivering, fatigue, dry cough, anorexia, anosmia, ageusia, dizziness, and sweating with COVID- 19. We also observed an association between higher mean age and abnormal CRP in COVID-19 patients. Interestingly, O- BG showed a protective effect against the affliction of COVID-19. Also, we showed a relationship between older age, history of heart disease and cancer and COVID-19 related mortality. Development of symptoms such as anosmia, dry cough, ageusia, fever and anorexia are also predictors of mortality of COVID-19 cases. O+BG is a protective factor against COVID-19 related mortality.

In our study, fever is the most significantly associated symptom with COVID-19, which is in line with Zhang et al. [19] and Chen et al. [20] findings. In contrast, Liang et al. [21], DeBiasi et al. [22], Tian et al. [23], Zho et al. [24], and Qin et al. [25] found a non-significant association between fever and COVID-19. Indeed, dyspnea is a relevant symptom with COVID-19 that is in parallel with Tian et al. [23], DeBiasi et al. [22] and Qin et al. [25] but is in contrast with the findings of Liang et al. [21] and Yan et al.[26]. Dizziness is associated with COVID-19 in our study, which is similar to the findings of Zhou et al. [27] and Zhang et al. [28]. But Shi et al. [17], Chen et al. [20] and Liang et al. [21] found no association with this. We did not find study regarding sweating, and to best of our knowledge, this is the first work to report this. Nikpouraghdam et al. [29] suggested an association between weakness and COVID-19 that is the same with our data but is in contrast with the findings of Shi et al. [17]. Shivering is observed in the cohort of COVID-19, which is reported by Zhu et al. [30] too. Fatigue is significantly related with COVID-19 in our cohort which confirms with the findings of Qin et al. [25] but is in contrast to the findings of Liang et al. [21], Tian et al. [23], Lei et al. [31], and Yan et al. [32]. A dry cough is reported by Du et al. [33] to be related with COVID-19, which is similar with our finding, while such an association is not found by Qin et al. [25], Lei et al. [31] and Shi et al. [17]. Anorexia is found to be a significant symptom for COVID- in our study. The association between anorexia and COVID-19 in our study is similar to Zhang et al. [19] and Chen et al. [20] but is not in line with the findings of Shi et al. [17], Lei et al. [31] and Qin et al. [25]. Anosmia is associated with COVID-19 in our cohort, which confirms with the findings of Yan et al. [26], Bagheri et al. [34] and Lee et al. [35]. Yan et al. [26] and Lee et al. [35] also introduced ageusia as an associated symptom with COVID-19 that is in line with our findings.

Regarding the association between age and COVID-19, we observed old age, as an indicator which is in line with Fan et al. [36], but is in contrast with the findings of Fu et al. [37], Fang et al. [38] and Omrani- Nava et al. [39], who did not find a significant association between age and COVID-19. This discrepancy could be explained by different demographic features (symptoms). We observed elevated CRP as a related feature with COVID-19 in our investigation that is parallel with Fu et al. [37] and is in contrast with the findings of Omrani-Nava et al. [39]. BCG vaccination showed no significant relationship between COVID- 19 versus. Healthy subjects in our cohort that is parallel with the findings of Li et al. [40] and Hamiel et al. [10] and in contrast to Dayal et al. [41].

Currently, there are additional data regarding predictor features of mortality related to COVID-19. Interestingly, some features with no obvious relationship with COVID-19 infection are associated with COVID-19 related mortality. O+BG, history of heart disease, and cancer are with COVID-19 infection showed a significant relationship with COVID-19 related mortality. While the development of symptoms such as anosmia, dry cough, ageusia, fever, and anorexia are the predictors of both COVID-19 infection and mortality. Old age is a predictor of mortality in Zhou et al. [24] report, that is also observed in our finding. Zhou et al. [24] reported other predictors of mortality such as fever, fatigue, myalgia, nausea, vomiting, diarrhea, cough and sputum that are not mortality predictors in our cohort. Fever and dyspnea are also considered as predictors of mortality by Iftime et al. [42] and Chen et al. [43], respectively, while we did not find an association between these symptoms with COVID-19 related mortality.

Interestingly, we observed more features associated with mortality in our cohort, but age seems to be the most important risk factor for COVID-19, and its induced mortality confirms with other published works [24, 42-45]. The predictive role of age is also confirmed by Sun et al., De Smet et al. [45] and Chen et al. [43]. In the old age, immune impairment can happen and also there is a possibility of respiratory disease. As mentioned above, comorbidities such as cancer and underlying heart diseases are associated with COVID-19 mortality. Iftimie et al. [42] observed such an association with type 2 diabetes mellitus (DM) and cancer, but they negated the relationship between the history of heart disease and COVID-19 related mortality. Regarding comorbidities, Ruan et al. [46] and Chen et al. [43] demonstrated the history of heart disease and COVID-19 related mortality. Indeed, a history of cerebrovascular disease is also determinant in Chen et al. [43] cohort. Male gender is associated with COVID-19 related mortality in Li et al. [47] study, but we did not observe such an association in our cohort. Positive CRP is a predictor in Ruan et al. [46] study, while it did not have any association with mortality in our investigation.

The association between certain type of blood group and COVID-19 related mortality is the novel findings of our investigation. We observed a protective role for O-BG against COVID-19 in our cohort that is similar to the findings of Zietz et al. [48] and Guo et al. [49]. Currently, there is no clear reason for such an association between certain BG and risk of COVID-19, but it has been suggested that O-BG or in other words, lack of A or B antigens might not permit viral entrance to cells with a subsequent lower chance for carriers to get infected by this virus. Guillon et al. [50] specifically inhibited adhesion of SARS-CoV S protein-expressing cells into ACE2-expressing cell lines by anti-A antibodies. Interestingly, we demonstrated a lower chance of mortality for O+BG carriers and further study is needed to support this. Summary of common findings reported by other state-of-the-art studies is listed in appendix tables 3 and 4. In appendix table 3, the effective features on COVID-19 infection are listed while in appendix Table 4, the effective features on the mortality rate of COVID-19 patients are listed.

**Table 3:**
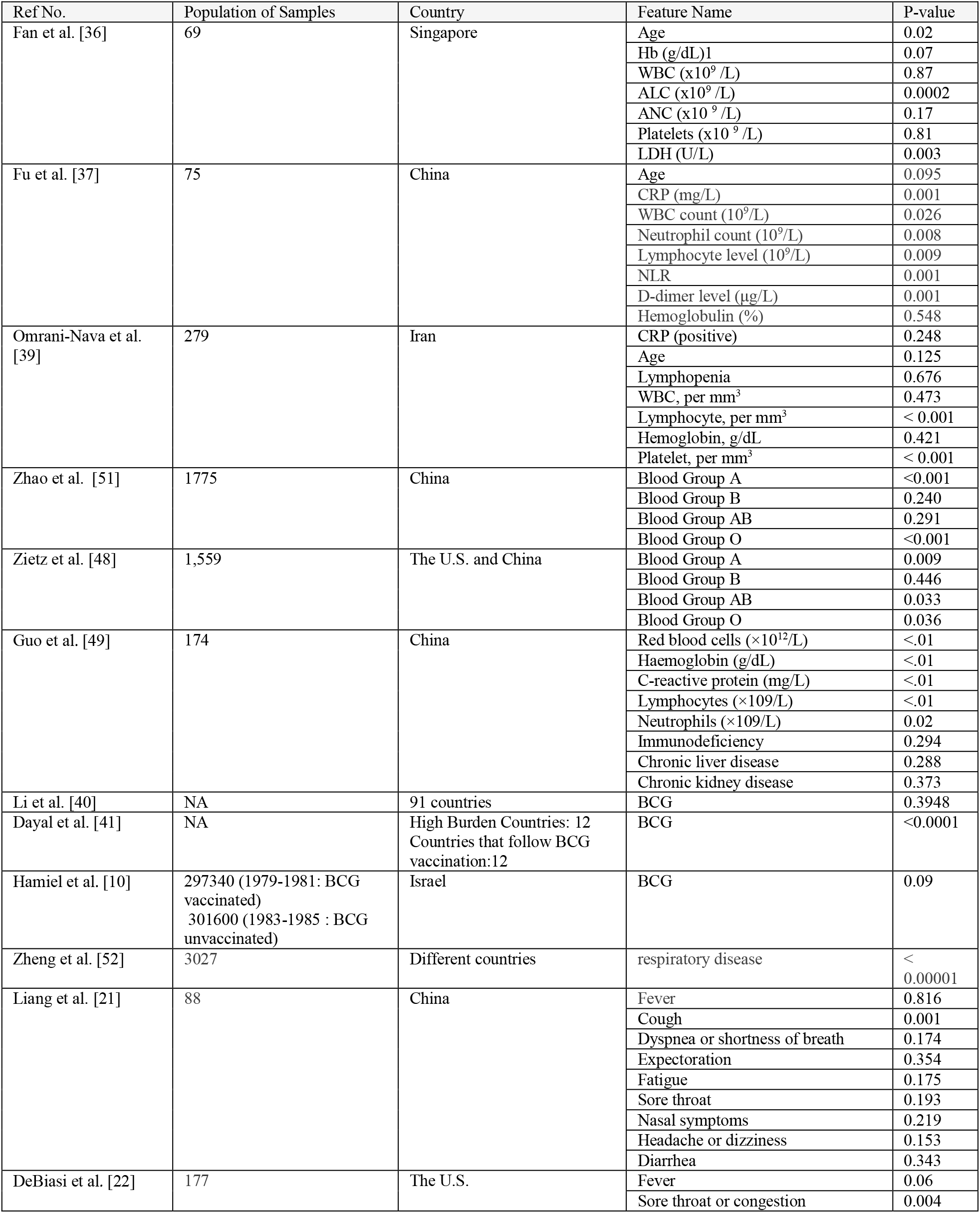
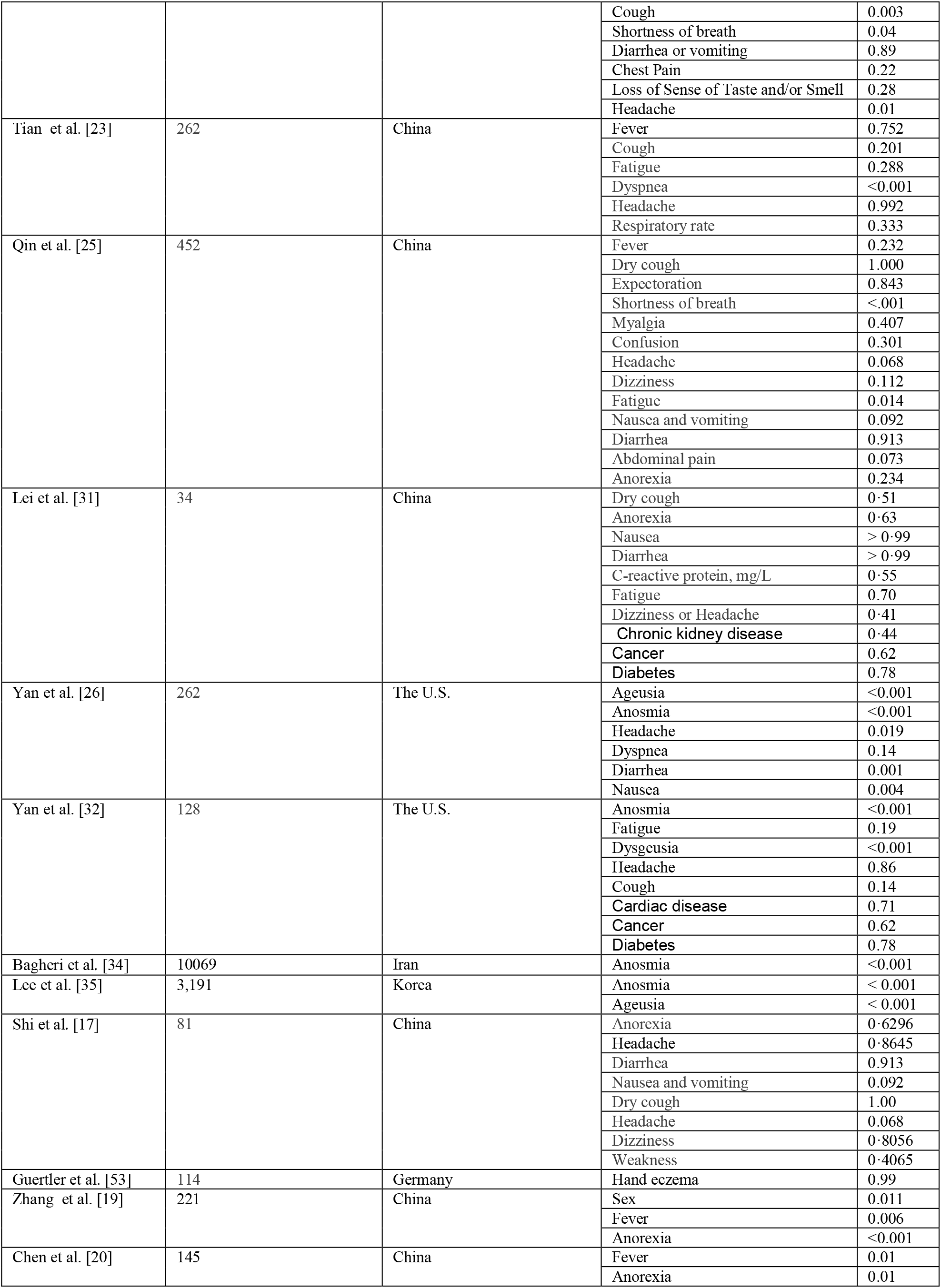
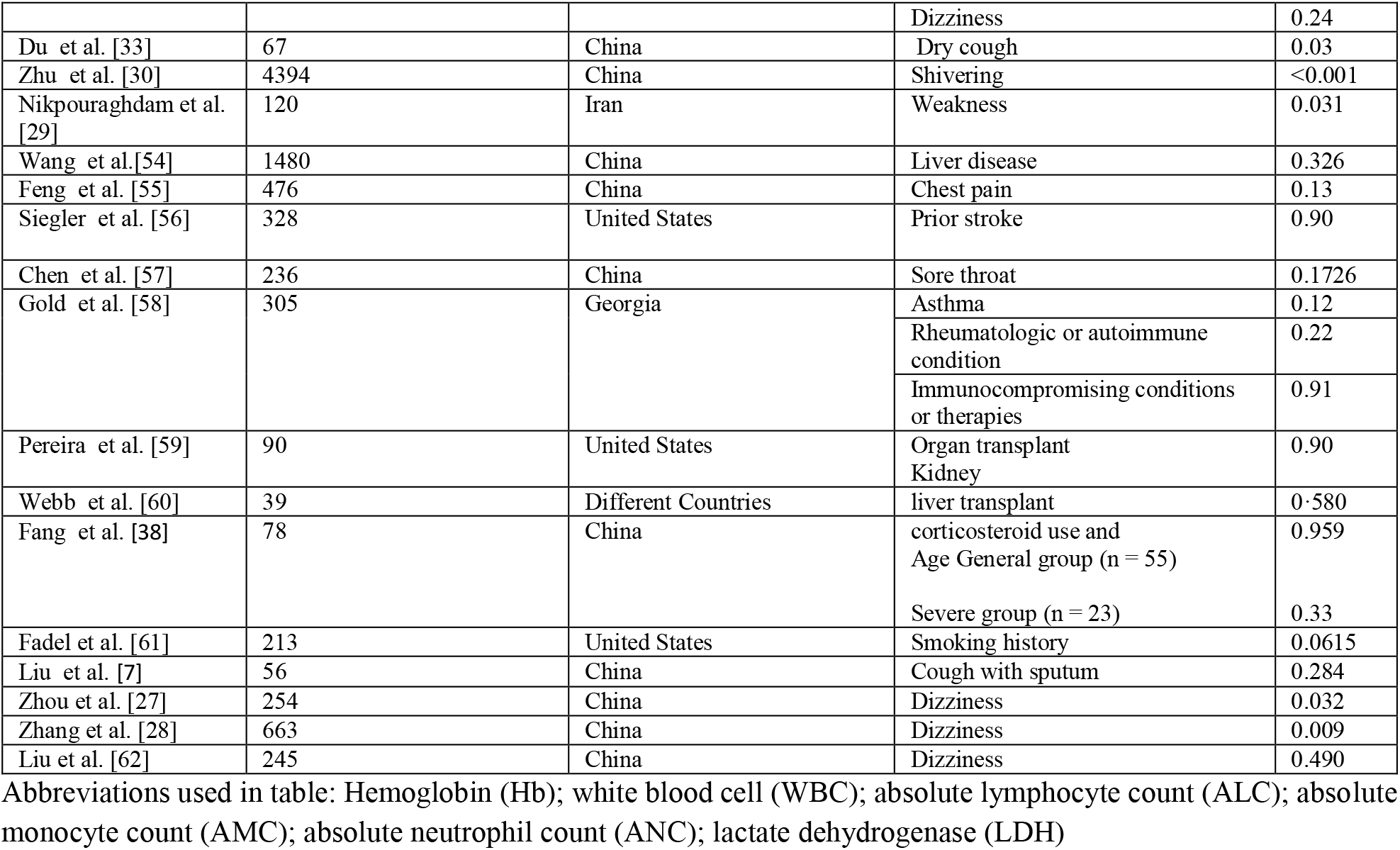
Summary of common findings reported by other state-of-the-art studies.

**Table 4.**
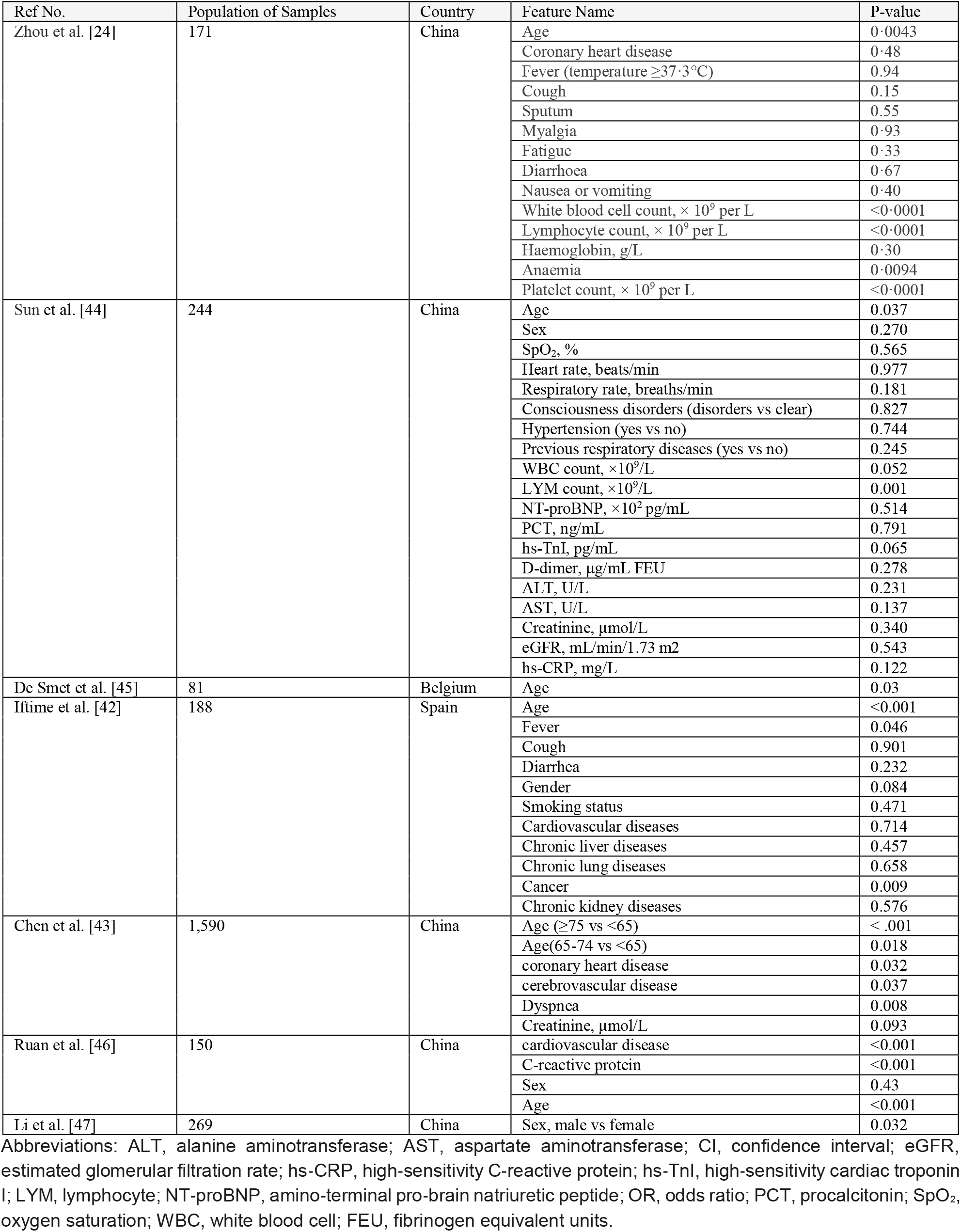
Some of the common clinical characteristics of COVID-19 patients with fatality.

The novelty of our study is that, we have investigated the predictive role of many reported features (symptoms) for both COVID-19 and its related mortality using 319 Iranian cases. This may give a clear trend of the whole population since the enrolled cases are selected from different regions of Iran. Indeed, data of mortality predictors are not yet widely available, especially in Middle East countries. Another point is that a combination of these features can help to manage the patient better and the need for upgrading the features associated with COVI-19 related mortality is essential in this pandemic era.

In conclusion, this is one of the first researches that investigate the effect of risk factors in prediction, clinical outcomes and mortality of COVID-19. Our results indicate tht, risk factors like fever, dyspnea, weakness, shivering and CRP are the most important factors in predicting the disease while age, O+BG, heart disease, anosmia and dry cough are the most crucial factors in the mortality of patients. This study may prove helpful in early prediction and risk reduction of mortality in patients infected with COVID-19. Further studies with more longitudinal follow-ups are needed to confirm our findings.

## Data Availability

The data are private.

## Appendix

## Notes

### Competing Interest Statement

The authors have declared no competing interest.

### Author Declarations

The study was approved by the Omid Hospital Ethics Committee.

